# Impact of COVID-19 vaccination on long-Term patient and kidney allograft survival following SARS-CoV-2 infection

**DOI:** 10.64898/2026.01.22.26344293

**Authors:** Ivana Dedinská, Darshana M. Dadhania, Carol Li, Nataliya Hauser, Perola Lamba, John R. Lee, Thangamani Muthukumar, Manikkam Suthanthiran

**Affiliations:** Transplant-Nephrology Department and Internal Medicine Department I, Jessenius Faculty of Medicine, Comenius University and University Hospital Martin, Slovakia; Division of Nephrology and Hypertension, Department of Medicine, New York-Presbyterian-Weill Cornell Medicine, New York, NY; Department of Transplantation Medicine, New York-Presbyterian Hospital-Weill Cornell Medicine, New York, NY

**Keywords:** kidney transplantation, SARS-CoV-2, COVID-19 vaccination, graft survival, acute kidney injury

## Abstract

The long-term impact of SARS-CoV-2 infection on kidney allograft survival remains incompletely understood, particularly regarding the influence of vaccination, acute kidney injury (AKI), and post-infection immunosuppression. We conducted a retrospective analysis of 129 kidney transplant recipients with confirmed SARS-CoV-2 infection between March 2020 and March 2022 with a median follow-up of 50 months. Among 129 recipients, 106 (82%) received vaccination at any time before or after SARS-CoV-2 infection (82%) while 23 (18%) remained unvaccinated. Unvaccinated patients experienced significantly lower long-term graft survival (52% vs. 85%; p = 0.0004) and patient survival (83% vs. 99%; p = 0.0003) compared with vaccinated recipients. AKI occurred in 15% of recipients and independently predicted graft failure (aHR 2.88; p = 0.0341). Post–SARS-CoV-2 serum creatinine and albuminuria were strong prognostic markers of graft loss. Unvaccinated status independently predicted graft failure in both transplantation-anchored (aHR 2.80; p = 0.0342) and SARS-CoV-2–anchored models (aHR 5.31; p = 0.0004). Continuation of mycophenolate mofetil at post-infection assessment was associated with reduced graft-failure risk (aHR 0.99; p = 0.0193). These findings underscore the importance of sustained vaccination in preserving long-term allograft function.

## 1 INTRODUCTION

Coronavirus disease 2019 (COVID-19), caused by severe acute respiratory syndrome coronavirus 2 (SARS-CoV-2), represents the most consequential global infectious disease outbreak of the past century. Kidney transplant recipients are among the most vulnerable groups due to their high comorbidity burden, lifelong immunosuppression, and limited ability to mount protective antiviral immune responses [1–4]. Since the onset of the pandemic, COVID-19 has been associated with excess mortality and acute kidney injury (AKI) in patients with chronic kidney disease, as well as frequent destabilization of allograft function in transplant recipients [5–8]. Pre-vaccination case-fatality rates exceeded 20–30%, particularly among hospitalized recipients [6,7].

Beyond the acute phase, accumulating evidence suggests that SARS-CoV-2 infection may exert long-term deleterious effects on kidney allografts through mechanisms including viral tropism for renal tissue, systemic immune activation, endothelial injury, complement activation, and microvascular thrombosis [9–13]. Emerging data indicate that SARS-CoV-2 infection in kidney transplant recipients may accelerate the decline in eGFR, particularly following severe COVID-19 episodes [14,15]. During acute infection, reduction or temporary withdrawal of mycophenolate mofetil was widely adopted early in the pandemic to enhance antiviral immunity [4,7,16,17]. However, the long-term consequences of antimetabolite modification on persistent graft injury and graft failure remain incompletely defined.

The introduction of SARS-CoV-2 vaccination fundamentally altered the clinical course of COVID-19 in the general population, markedly reducing the risk of severe disease, hospitalization, and death [18–20]. In transplant recipients, however, vaccine immunogenicity is impaired, with attenuated seroconversion rates, reduced neutralizing antibody titers, and waning cellular immunity over time [21–25].

Most studies evaluating vaccination in kidney transplant recipients have focused predominantly on short-term outcomes, with follow-up rarely exceeding 6–12 months [26–33]. In contrast, data on long-term allograft outcomes after vaccination remain sparse and largely limited to short-term assessments of rejection or graft dysfunction [34]. Consequently, whether early survival benefits translate into durable protection against progressive graft injury and late graft failure remains insufficiently characterized. While traditional transplant risk factors such as donor-specific antibodies, delayed graft function, and immunologic sensitization shape baseline allograft vulnerability [35–37], the impact of maintenance immunosuppression after SARS-CoV-2 infection, particularly the continuation or modification of antimetabolite therapy, has not been adequately defined.

Accordingly, we conducted a comprehensive long-term retrospective study of kidney transplant recipients with documented SARS-CoV-2 infection. The primary objective was to determine whether COVID-19 vaccination status is associated with long-term graft and patient survival. Secondary objectives were: (i) to assess the impact of AKI and hospitalization during COVID-19 on subsequent graft survival; (ii) to evaluate whether post-infection allograft function predicts long-term graft failure; and (iii) to identify independent predictors of graft loss using both transplantation-anchored and SARS-CoV-2–anchored multivariable Cox regression models.

## 2 MATERIAL AND METHODS

### 2.1 Study Conduct

This retrospective cohort study was conducted and reported in accordance with the Strengthening the Reporting of Observational Studies in Epidemiology (STROBE) guidelines. We have reported our initial observation of the impact of SARS-CoV-2 infection on 90-day mortality and graft loss in 54 kidney transplant recipients during the initial phase of the pandemic [1,2].

For the present analysis, we included kidney transplant recipients transplanted at Weill Cornell – New York Presbyterian Hospital and who had laboratory-confirmed SARS-CoV-2 infection and consented to participate in the longitudinal observational study entitled “Impact of Covid-19 illness on kidney transplant candidates and recipients” (IRB protocol 20-05022154). The study protocol was approved by WCM Institutional Review Board and adhered to the ethical principles outlined in The Declaration of Helsinki.

The present study represents the first-long term, longitudinal investigation of its kind, encompassing 129 patients, and reporting long-term patient and kidney graft survival. In addition, we identify adverse outcomes in the subgroup of recipients who remained unvaccinated against SARS-CoV-2.

### 2.2 Study Design and Population

Potential participants were identified through retrospective chart review and clinician referral during the recruitment phase. The inclusion criteria were: (1) Adult kidney transplant recipients transplanted at WCM-NYP, (2) laboratory-confirmed SARS-CoV-2 infection by polymerase chain reaction or antigen testing between March 20, 2020, and March 7, 2022, (3) ability to provide informed consent for prospective monitoring of clinical parameters. The exclusion criteria were (1) Lack of laboratory confirmation of SARS-CoV-2 infection and (2) absence of longitudinal follow-up at WCM-NYP.

A retrospective chart review of 132 kidney transplant recipients was conducted to obtain all transplant-related and SARS-CoV-2 infection-related variables and 129 individuals met the inclusion and exclusion criteria (Figure 1). Data on kidney function, immunosuppressive therapies, and rejection were collected at enrollment and monitored prospectively through September 2025. Median time from transplant to SARS-CoV-2 infection was 48 months, median time from SARS-CoV-2 infection to enrollment was 8 months (239 days) and median follow-up post SARS-CoV-2 infection was 50 months.

**Figure 1.**
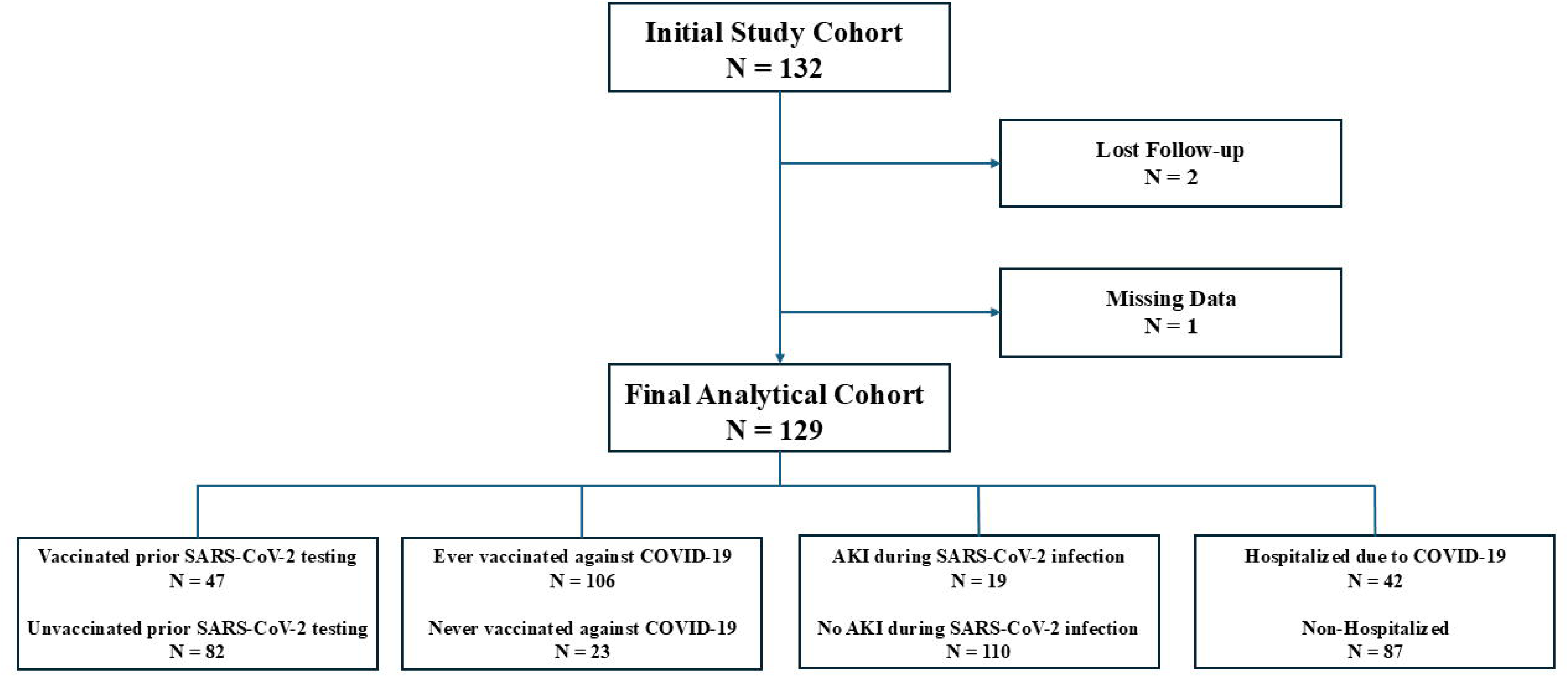
Study flow diagram of kidney transplant recipients with confirmed SARS-CoV-2 infection. A total of 132 kidney transplant recipients with laboratory-confirmed SARS-CoV-2 infection were initially identified. Two patients were excluded due to loss to follow-up and one patient due to missing key clinical data. The final analytical cohort comprised 129 patients. The cohort was further stratified according to vaccination status prior to SARS-CoV-2 testing (vaccinated vs. unvaccinated), ever vaccination status (ever vaccinated vs. never vaccinated), occurrence of acute kidney injury (AKI) during SARS-CoV-2 infection (AKI vs. no AKI), and hospitalization due to COVID-19 (hospitalized vs. non-hospitalized).

### 2.3 Clinical Data Collection

Baseline demographic, clinical, donor, and transplant-related characteristics were extracted from the EPIC electronic health record system. SARS-CoV-2 infection was classified as asymptomatic or symptomatic. Hospitalization was defined as admission primarily for management of COVID-19–related complications. Acute kidney injury during SARS-CoV-2 infection was defined as an absolute increase in serum creatinine of at least 0.5 mg/dL from baseline, or a relative increase of at least 30%, or the initiation of renal replacement therapy. Vaccination status was categorized as ever vaccinated, defined as receipt of any COVID-19 vaccine before or after SARS-CoV-2 infection, based on review of medical records through September 2025. Changes in maintenance immunosuppression during SARS-CoV-2 infection were systematically assessed, including discontinuation, dose reduction, or no modification of tacrolimus and mycophenolate mofetil (MMF). Analyses were restricted to recipients receiving the respective agent prior to infection. Maintenance immunosuppression at enrollment was recorded, including tacrolimus trough levels, daily MMF dose, and corticosteroid use.

When available, results of the most recent clinically indicated kidney biopsy and donor-specific antibody (DSA) testing performed before and after SARS-CoV-2 infection were extracted. Additional methodological details, including complete variable definitions and data collection procedures, are provided in Supplementary Methods 1A.

### 2.4 Follow-Up

Longitudinal kidney graft function was tracked using a REDCap database.. Long-term outcome measures included (1) all-cause mortality and (2) kidney graft loss, defined as return to dialysis or re-transplantation.

### 2.5 Ethical approval

All procedures involving human participants were approved by the institutional research committee and adhered to the ethical standards of the 1964 Helsinki Declaration and its later amendments.

Clinical and research activities complied with the Principles of the Declaration of Istanbul as outlined in the Declaration of Istanbul on Organ Trafficking and Transplant Tourism.

### 2.6 Statistical analysis

Predefined group comparisons included: (i) ever vaccinated versus never vaccinated, (ii) the three vaccination categories (vaccinated prior to SARS-CoV-2 testing, vaccinated after testing, never vaccinated), (iii) AKI associated with SARS-CoV-2 infection versus no AKI, and (iv) hospitalized versus non-hospitalized patients with COVID-19. A two-sided p value <0.05 was considered statistically significant. All analyses were performed on complete cases.

Statistical analyses were performed using MedCalc version 23.3.7 (Ostend, Belgium). Continuous variables are presented as mean ± standard deviation or median (interquartile range), as appropriate, and were compared using the Student’s t test or Mann–Whitney U test (two groups) and one-way ANOVA or Kruskal–Wallis test (three groups). Categorical variables are presented as number (percentage) and were compared using the chi-square test or Fisher’s exact test, as appropriate.

Graft and patient survival were analyzed using Kaplan–Meier methods within transplantation-anchored and post–SARS-CoV-2 infection–anchored time-to-event frameworks; survival curves were compared using the log-rank test. Time zero was defined as either the date of kidney transplantation or the date of the first positive SARS-CoV-2 test.

Univariable and multivariable logistic regression analyses were used to identify factors associated with AKI and hospitalization due to COVID-19. Results are reported as odds ratios (ORs) with 95% confidence intervals (CIs).

Univariable and multivariable Cox proportional hazards regression was used to evaluate predictors of post–SARS-CoV-2 graft loss and long-term transplantation-anchored graft survival using prespecified covariate-adjusted models. Results are reported as hazard ratios (HRs) with 95% confidence intervals (CIs).

Detailed description of time origins, covariate selection, model construction, and handling of collinearity are provided in Supplementary Statistical Methods (Supplementary Methods 1B).

## 3 RESULTS

The median age at time of kidney transplantation among 129 kidney transplant recipients with confirmed SARS-CoV-2 infection who met the inclusion and exclusion criteria was 52 years (IQR 42–60). The cohort was 57% male, with a racial/ethnic distribution of 38% white, 27% black, and 25% Hispanic (Supplementary Table S1). Most patients (66%) were receiving dialysis prior to transplantation, while 31% underwent preemptive transplantation. Living-donor transplantation accounted for 58% of cases. Among the deceased-donor kidney recipients, 43% experienced delayed graft function.

The median time from transplantation to SARS-CoV-2 infection was 48 months (IQR 9.75-101.5). COVID-19–related hospitalization occurred in 33% of patients, and 15% developed acute kidney injury. At last follow-up in September 2025, patient survival was 96%, and graft survival was 79% following SARS-CoV-2 infection. Detailed recipient, transplant, virological, clinical, and laboratory characteristics are provided in Supplementary Tables S1–S5 and Supplementary Figures S1–S5.

### 3.1 Impact of COVID-19 Vaccination Status

Among the 129 kidney transplant recipients, 106 (82%) received at least one dose of a COVID-19 vaccine during follow-up (“ever vaccinated”), while 23 (18%) remained unvaccinated throughout the observation period (“never vaccinated”). Vaccination status was documented in the health records by the care providers. Baseline demographic characteristics, comorbidities, transplant-related factors, and immunologic risk profile - including age at transplantation, sex, race, sensitization status and early graft function were comparable between the two groups (Table 1).

**Table 1.**
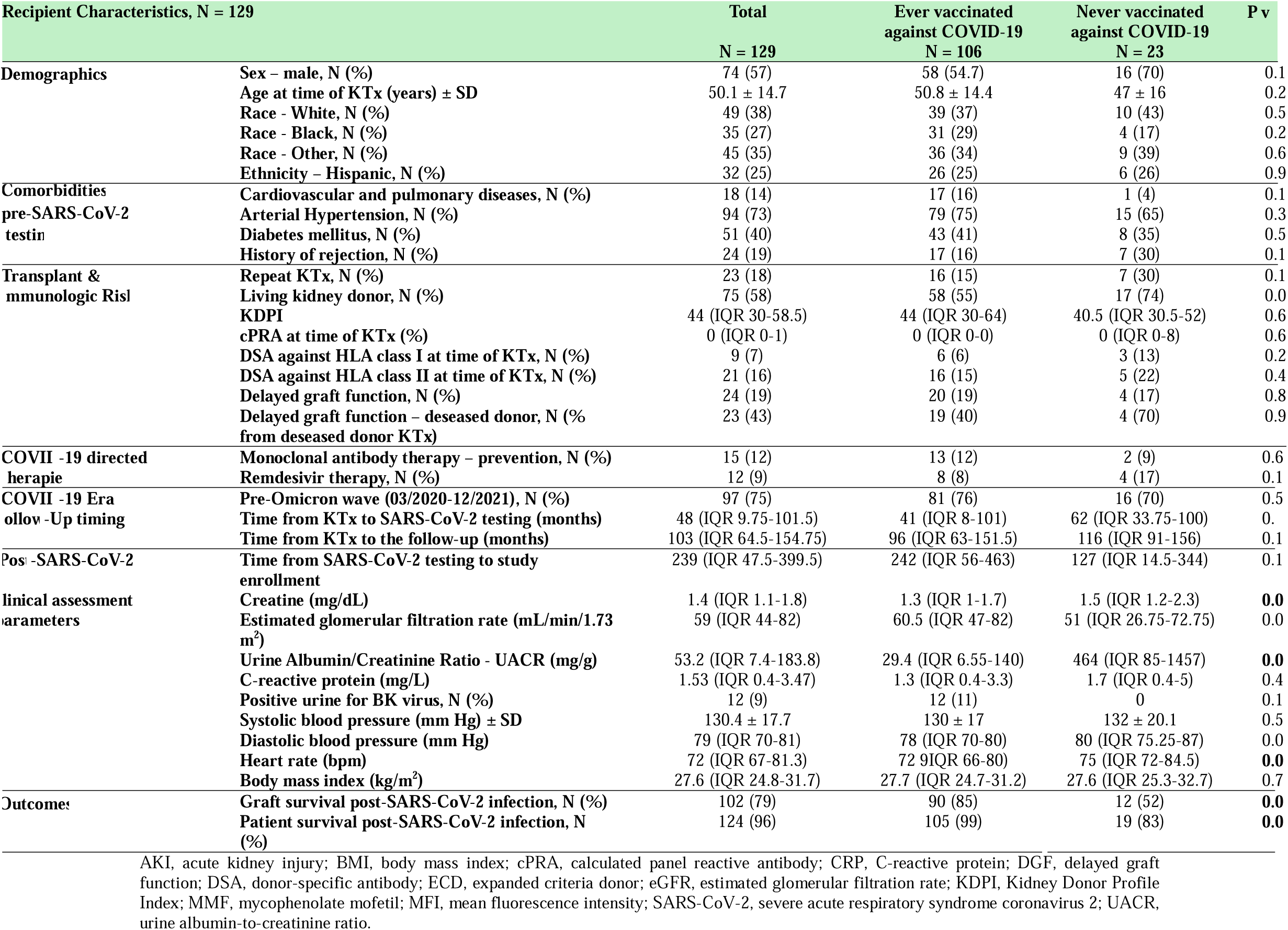
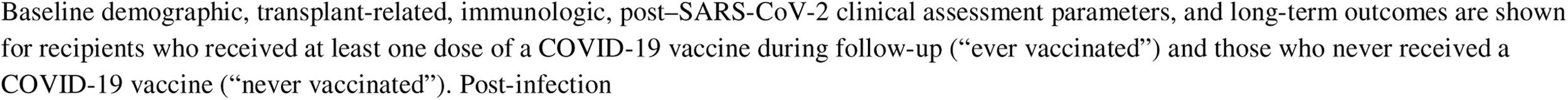
Characteristics and long-term outcomes of kidney transplant recipients according to ever versus never COVID-19 vaccination status.

COVID-19 vaccines became publicly available in New York City began in March 2021. Because 42% of ever-vaccinated and 43% of never-vaccinated recipients were infected before vaccines became available (p = 0.9301), suggesting limited access does not explain the poorer outcomes among never-vaccinated patients (Supplementary Figure S6). The distribution of COVID-19 vaccine types is shown in Supplementary Figure S7.

Clinical characteristics and outcomes differed significantly between vaccinated and unvaccinated recipients. Never-vaccinated recipients exhibited higher serum creatinine at post-infection assessment (median 1.5 vs. 1.3 mg/dL, p = 0.0298) and markedly elevated albuminuria (median urine albumin-creatinine ratio - UACR 464 vs. 29 mg/g, p = 0.0001) compared with ever-vaccinated patients. Estimated glomerular filtration rate (eGFR) was lower in never-vaccinated recipients, although this difference did not reach statistical significance.

Most importantly, vaccination status was strongly associated with long-term graft and patient survival. Graft survival was significantly lower among never-vaccinated recipient compared with ever-vaccinated recipients (52% vs. 85%, p = 0.0004), and patient survival was likewise significantly reduced (83% vs. 99%, p = 0.0003) (Table 1).

Kaplan–Meier survival curves of kidney graft and patient survival, anchored to both the time of kidney transplantation and to the time of SARS-CoV-2 infection, are shown in Figure 2A-2B and Figure 2C-2D, respectively. Never-vaccinated recipients experienced a significantly inferior long-term patient survival and graft survival compared with ever-vaccinated recipients.

**Figure 2.**
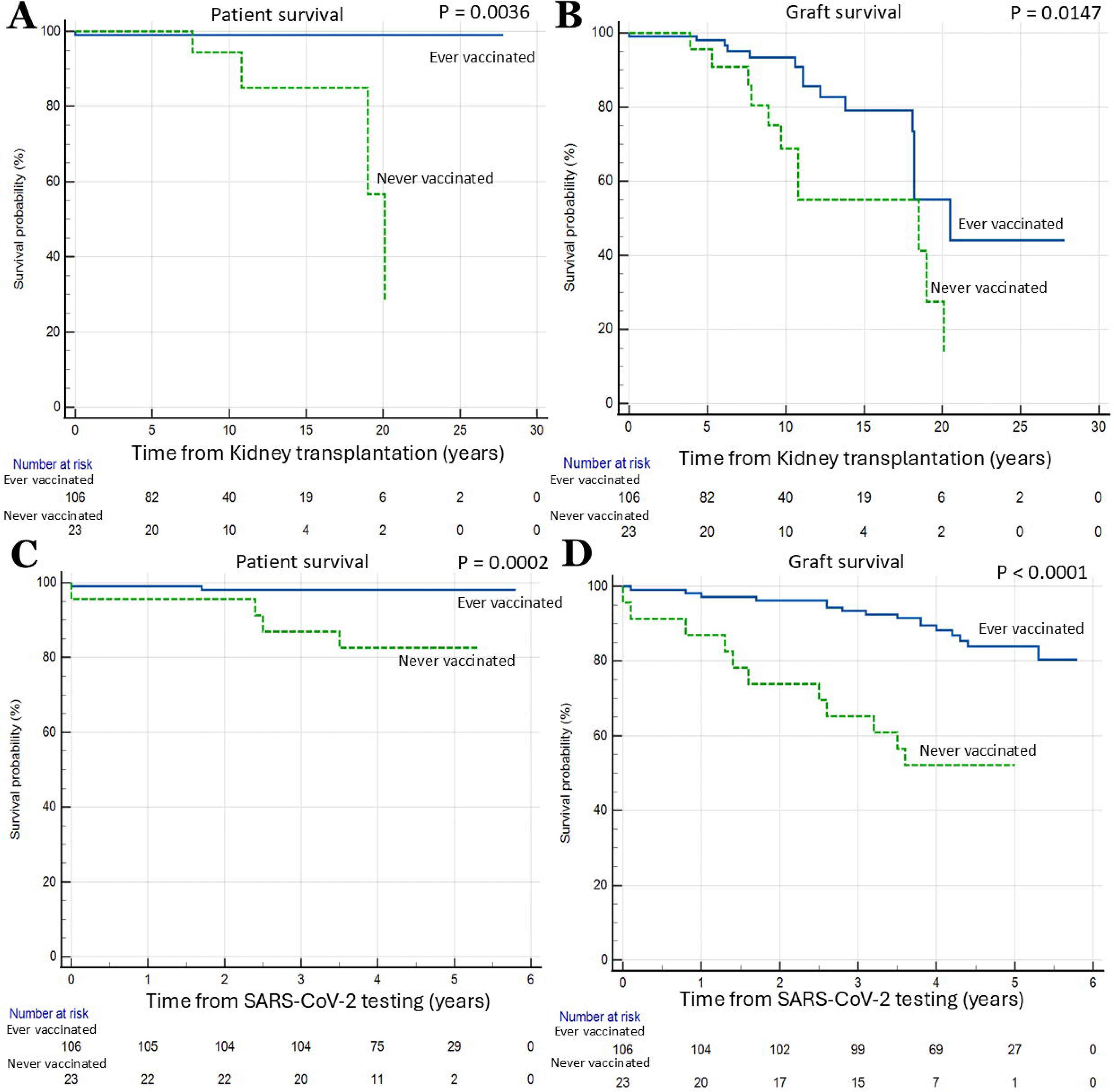
Transplantation-anchored and Post–SARS-CoV-2 infection patient and graft survival according to ever COVID-19 vaccination status. (A) Kaplan–Meier analysis of long-term patient survival in ever-vaccinated versus never-vaccinated kidney transplant recipients (log-rank p = 0.0036). (B) Kaplan–Meier analysis of long-term graft survival in ever-vaccinated versus never-vaccinated recipients (log-rank p = 0.0147). (C) Kaplan–Meier analysis of post-infection patient survival in ever-vaccinated versus never-vaccinated recipients (log-rank p = 0.0002). (D) Kaplan–Meier analysis of post-infection graft survival in ever vaccinated versus never-vaccinated kidney transplant recipients (log-rank p < 0.0001). Time zero was defined as the date of kidney transplantation (A, B). Time zero was defined as the date of the positive SARS-CoV-2 test (C, D). Survival curves were compared using the log-rank test. Numbers at risk are shown below each panel. Blue solid lines indicate ever-vaccinated recipients, and green dashed lines indicate never-vaccinated recipients. *P* values are displayed in each panel.

In contrast, vaccination status specifically at the time of SARS-CoV-2 infection (vaccinated prior to testing vs unvaccinated at testing) was not associated with significant differences in long-term graft survival or patient survival (Supplementary Figures S8).

Analysis of three vaccination categories showed a uniform pattern of poorer outcomes among never-vaccinated recipients (Supplementary Table S6). Across all four Kaplan–Meier analyses—transplantation-anchored patient survival, transplantation-anchored graft survival, SARS-CoV-2–anchored patient survival, and SARS-CoV-2–anchored graft survival—never-vaccinated recipients experienced significantly worse survival (Supplementary Figure S9).

### 3.2 Acute Kidney Injury Associated with SARS-CoV-2 infection

Among the 129 kidney transplant recipients, 19 patients (15%) developed acute kidney injury (AKI) during SARS-CoV-2 infection, while 110 patients (85%) did not. Baseline demographic, transplant-related, immunologic, and clinical characteristics stratified by AKI status are shown in Supplementary Table S7. All patients who developed AKI required hospitalization during the acute infection.

#### Predictors of AKI associated with SARS-CoV-2 infection

Univariable and multivariable logistic regression analyses were performed to identify factors associated with the development of AKI associated with SARS-CoV-2 infection (Table 2). In univariable analyses, clinical features indicative of greater acute illness severity-dyspnea and diarrhea-were strongly associated with AKI. Additional risk factors, included retransplant status (≥ second kidney transplant), unvaccinated status, and absence of anti–SARS-CoV-2 spike antibodies prior to infection.

**Table 2.** Univariable and multivariable logistic regression analysis of factors associated with acute kidney injury associated with SARS-CoV-2 infection.

| Outcome: Acute Kidney Injury during COVID-19 | Unadjusted OR (95% CI) | p-value | Adjusted OR (95% CI) | p-value |
| --- | --- | --- | --- | --- |
| Age at time of KTx (years) | 0.97 (0.94-1.00) | 0.1500 | 0.97 (0.93-1.01) | 0.1517 |
| Diabetes mellitus | 1.13 (0.42-3.04) | 0.8041 | 1.56 (0.455-5.39) | 0.4762 |
| Time from KTx to SARS-CoV-2 testing (months) | 1.0014 (0.9936-1.0093) | 0.7178 | 0.9998 (0.9912-1.0085) | 0.9649 |
| Sex – male | 0.36 (0.13-1.00) | 0.0506 | 0.38 (0.13-1.12) | 0.0815 |
| Race - Black | 1.27 (0.44-3.66) | 0.6541 | 1.32 (0.40-4.29) | 0.6436 |
| Ethnicity – Hispanic | 0.51 (0.14-1.90) | 0.3218 | 0.45 (0.10-1.87) | 0.2741 |
| History of rejection | 2.35 (0.79-7.02) | 0.1232 | 2.20 (0.66-7.25) | 0.1947 |
| Repeat KTx | 3.42 (1.17-10.01) | **0.0244** | 3.29 (0.67-15.99) | 0.1392 |
| Deceased donor | 2.14 (0.79-5.75) | 0.1306 | 1.36 (0.38-4.89) | 0.6289 |
| KDPI | 0.98 (0.95-1.02) | 0.5600 | 1.01 (0.96-1.07) | 0.5794 |
| cPRA at time of KTx | 1.01 (0.99-1.02) | 0.1412 | 1.01 (0.99-1.03) | 0.1973 |
| DSA at time of KTx | 3.46 (1.16-10.26) | **0.0252** | 3.28 (0.95-11.32) | 0.0594 |
| Delayed graft function | 1.18 (0.35-3.95) | 0.7807 | 1.30 (0.35-4.78) | 0.6836 |
| DSA before COVID-19 | 0.98 (0.28-3.40) | 0.9819 | 0.90 (0.22-3.72) | 0.8929 |
| Symptoms at time of infection |  |  |  |  |
| - Fever | 1.37 (0.50-3.76) | 0.5327 | 1.40 (0.47-4.09) | 0.5375 |
| - Cough | 1.60 (0.49-5.20) | 0.4291 | 1.58 (0.44-5.68) | 0.4776 |
| - Fatigue or Myalgia | 1.37 (0.51-3.67) | 0.5261 | 1.55 (0.53-4.54) | 0.4224 |
| - Diarrhea | 5.32 (1.90-14.86) | **0.0014** | 7.55 (2.19-26.04) | **0.0014** |
| - Short of breath | 7.00 (2.41-20.25) | **0.0003** | 9.84 (2.86-33.80) | **0.0003** |
| - Loss of taste and smell | 0.40 (0.08-1.85) | 0.2409 | 0.37 (0.07-1.83) | 0.2254 |
| - Headaches | 1.09 (0.36-3.33) | 0.8690 | 0.81 (0.22-2.97) | 0.7520 |
| - Confusion | 0.55 (0.06-4.61) | 0.5861 | 0.63 (0.06-5.79) | 0.6861 |
| Pre-Omicron wave (03/2020-12/2021) | 0.50 (0.17-1.41) | 0.1940 | 0.53 (0.17-1.62) | 0.2707 |
| Unvaccinated status | 3.42 (1.17-10.01) | **0.0244** | 4.95 (1.43-17.06) | **0.0112** |
| Number of vaccine doses prior to infection | 0.99 (0.31-3.09) | 0.9902 | 1.16 (0.28-4.77) | 0.8303 |
| Months between last vaccine dose and COVID-19 | 0.73 (0.46-1.14) | 0.1685 | 0.72 (0.45-1.18) | 0.1998 |
| Positive anti-S* antibodies before COVID-19 | 0.12 (0.03-0.41) | **0.0007** | 0.04 (0.009-0.24) | **0.0003** |
| Monoclonal antibody therapy (any) | 0.87 (0.18-4.24) | 0.8712 | .57 (0.10-3.13) | 0.5234 |

\*anti-spike protein SARS-CoV-2 antibody
AKI, acute kidney injury; BMI, body mass index; CI, confidence interval; cPRA, calculated panel reactive antibody; CRP, C-reactive protein; DGF, delayed graft function; DSA, donor-specific antibody; eGFR, estimated glomerular filtration rate; KDPI, Kidney Donor Profile Index; MMF, mycophenolate mofetil; MFI, mean fluorescence intensity; OR, odds ratio; SARS-CoV-2, severe acute respiratory syndrome coronavirus 2; UACR, urine albumin-to-creatinine ratio
Univariable and multivariable logistic regression models were used to evaluate clinical, transplant-related, immunologic, and COVID-19-related factors associated with the development of acute kidney injury (AKI) during SARS-CoV-2 infection. AKI was analyzed as a binary outcome variable (yes/no).
Given the limited number of AKI events (n = 19), a parsimonious adjustment strategy was prespecified to avoid model overfitting. For each exposure of interest, a separate multivariable model was fitted that included the exposure together with the following baseline covariates: recipient age at transplantation, diabetes mellitus, and time from kidney transplantation to SARS-CoV-2 diagnosis. Sex was additionally included in sensitivity analyses and did not materially change the results.
Odds ratios (ORs) with 95% confidence intervals (CIs) are reported. p-value < 0.05 was considered statistically significant.

In multivariable models adjusted for recipient age at transplantation, diabetes mellitus, and time from kidney transplantation to SARS-CoV-2 diagnosis, dyspnea (OR 9.84, p = 0.0003) and diarrhea (OR 7.55, p = 0.0014) remained independently associated with the development of AKI. Unvaccinated status (OR 4.95, p = 0.0112) continued to confer substantially higher odds of AKI, whereas detectable anti–SARS-CoV-2 spike antibodies prior to infection were independently protective (OR 0.04, p = 0.0003).

#### Impact of AKI on Post–SARS-CoV-2 Graft and Patient Survival

Post–SARS-CoV-2 graft and patient survival were evaluated using Kaplan–Meier analyses anchored to the date of the positive SARS-CoV-2 diagnosis. Recipients who developed AKI during SARS-CoV-2 infection demonstrated significantly inferior post-infection graft survival compared with those without AKI. In contrast, post–SARS-CoV-2 patient survival did not differ significantly according to AKI status (Figures 3A and 3B).

**Figure 3.**
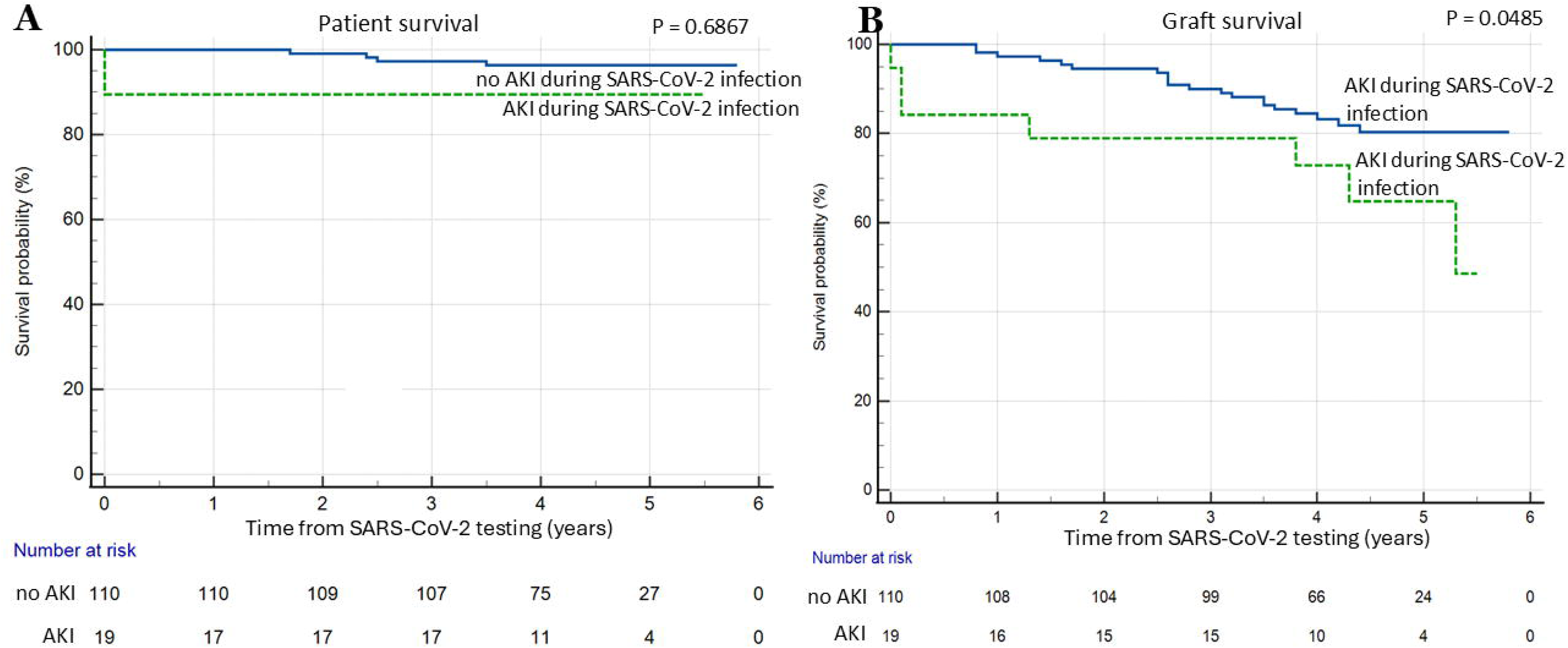
Post–SARS-CoV-2 patient and graft survival according to acute kidney injury associated with SARS-CoV-2 infection. (A) Kaplan–Meier analysis of post–SARS-CoV-2 patient survival according to AKI status during infection, showing no significant difference between groups (log-rank p = 0.6867). (B) Kaplan–Meier analysis of post–SARS-CoV-2 graft survival stratified by the presence of acute kidney injury (AKI) during SARS-CoV-2 infection (log-rank p = 0.0485). Time zero was defined as the date of the positive SARS-CoV-2 test. Numbers at risk are shown below each panel. Blue solid lines indicate recipients without acute kidney injury (AKI), and green dashed lines indicate recipients with AKI during SARS-CoV-2 infection. *P* values are displayed in each panel.

When survival time was measured from the date of kidney transplantation (Supplementary Figure S9), AKI showed a similar pattern, trend toward lower graft survival (log-rank p = 0.0668) but no significant difference in patient survival (log-rank p = 0.5039).

### 3.3 Hospitalization Due to COVID-19

Among 129 kidney transplant recipients with documented SARS-CoV-2 infection, 42 (32.6%) patients required hospitalization due to COVID-19, while 87 (67.4%) were managed in the outpatient setting. Baseline demographic, transplant-related, immunologic, and clinical characteristics according to hospitalization status are summarized in Supplementary Table S8.

Hospitalized patients had a significantly higher burden of cardiopulmonary comorbidities compared with non-hospitalized recipients (24% vs. 9%, p = 0.0215). AKI occurred exclusively in hospitalized patients (45% vs. 0%, p < 0.0001). Hospitalized recipients more frequently presented with fever, cough, dyspnea, and diarrhea, whereas asymptomatic infection occurred only among non-hospitalized patients (21%, p = 0.0014). Remdesivir treatment was administered exclusively to hospitalized recipients (29% vs. 0%, p < 0.0001).

Long-term graft survival (71% vs. 83%, p = 0.1177) and patient survival (93% vs. 98%, p = 0.1563) were numerically lower in hospitalized patients but did not reach statistical significance (Supplementary Table S8). Kaplan–Meier analyses of transplantation-anchored and post-infection graft and patient survival according to hospitalization status are shown in Supplementary Figure S10.

In logistic regression analyses, dyspnea (OR 23.15, p < 0.0001) was the strongest predictor of hospitalization, with diarrhea, fever, and cough also independently associated. Detectable anti–SARS-CoV-2 spike antibodies prior to infection was independently protective (Supplementary Table S9).

### 3.4 Independent Predictors of Graft Loss in Transplantation-Anchored and SARS-CoV-2–Anchored Cox Regression Models

#### SARS-CoV-2–Infection–Anchored Cox Regression (Model A and Model B)

To identify predictors of graft loss after SARS-CoV-2 infection, we performed Cox proportional hazards analyses anchored to the time of SARS-COV-2 infection using two complementary modeling frameworks.

Model A focused on acute COVID-19 course and pre-infection immune status. Unvaccinated status (aHR 5.31, p = 0.0004) and AKI during SARS-CoV-2 infection (aHR 2.88, p = 0.0341) were independently associated with subsequent graft loss after adjustment (Table 3).

**Table 3.** Unadjusted and adjusted SARS-CoV-2–anchored Cox proportional hazards regression analysis (Model A) of predictors of post–SARS-CoV-2 graft loss.

| Outcome: graft loss | Unadjusted HR (95% CI) | p-value | Adjusted HR (95% CI) | p-value |
| --- | --- | --- | --- | --- |
| Sex – male | 0.76 (0.35-1.61) | 0.4766 | 0.77 (0.35-1.71) | 0.5329 |
| Age at time of KTx (years) | 0.99 (0.97-1.02) | 0.9674 | 0.98 (0.95-1.02) | 0.5811 |
| Deceased donor | 1.27 (0.60-2.71) | 0.5256 | 1.27 (0.58-2.76) | 0.5349 |
| KDPI | 1.01 (0.98-1.05) | 0.2677 | 1.04 (0.99-1.09) | 0.1132 |
| History of rejection | 2.61 (1.17-5.84) | **0.0188** | 2.37 (0.93-6.02) | 0.0680 |
| Time from KTx to SARS-CoV-2 testing (months) | 1.00 (1.00-1.01) | **0.0065** | 1.01 (0.98-1.05) | 0.4581 |
| Symptoms at time of infection |  |  |  |  |
| - Hospitalization for COVID-19 | 1.76 (0.82-3.77) | 0.1455 | 1.30 (0.55-3.10) | 0.5410 |
| - AKI associated with COVID-19 | 2.31 (0.97-5.48) | 0.0558 | 2.88 (1.08-7.68) | **0.0341** |
| - Fever | 1.52 (0.68-3.40) | 0.2985 | 1.65 (0.67-4.03) | 0.2702 |
| - Cough | 1.10 (0.46-2.60) | 0.8236 | 1.16 (0.42-3.20) | 0.7617 |
| - Fatigue or Myalgia | 0.60 (0.27-1.30) | 0.1971 | 0.53 (0.23-1.21) | 0.1348 |
| - Diarrhea | 0.93 (0.37-2.31) | 0.8801 | 0.93 (0.34-2.56) | 0.9035 |
| - Short of breath | 0.92 (0.40-2.11) | 0.8482 | 0.79 (0.32-1.94) | 0.6226 |
| - Loss of taste and smell | 0.65 (0.22-1.90) | 0.4400 | 0.38 (0.10-1.42) | 0.1521 |
| - Headaches | 0.46 (0.16-1.35) | 0.1624 | 0.60 (0.19-1.86) | 0.3809 |
| - Confusion | 1.38 (0.41-4.61) | 0.5942 | 1.32 (0.37-4.69) | 0.6679 |
| Pre-Omicron wave (03/2020-12/2021) | 0.54 (0.22-1.29) | 0.1693 | 0.73 (0.28-1.87) | 0.5184 |
| Unvaccinated status | 4.89 (2.23-10.72) | **0.0001** | 5.31 (2.09-13.49) | **0.0004** |
| 3 ≥ vaccine doses prior to infection | 2.15 (0.25-18.02) | 0.4790 | 0.20 (0.11-12.56) | 0.8788 |
| 3 ≥ months between last vaccine dose and COVID | 1.05 (0.23-4.71) | 0.9429 | 1.05 (0.17-6.17) | 0.9542 |
| Positive anti-S* antibodies before COVID-19 | 0.62 (0.23-1.67) | 0.3540 | 0.65 (0.24-1.75) | 0.4006 |
| Remdesivir treatment | 4.20 (1.68-10.50) | **0.0021** | 1.82 (0.53-6.21) | 0.3364 |
| Monoclonal antibody therapy (any) | 1.13 (0.33-3.79) | 0.8430 | 1.41 (0.36-5.43) | 0.6118 |
| Tacrolimus dose reduced | 1.62 (0.64-4.08) | 0.3019 | 1.12 (0.43-2.88) | 0.8146 |
| MMF dose reduced during SARS-CoV-2 | 1.29 (0.50-3.32) | 0.5883 | 1.58 (0.62-4.04) | 0.3351 |
| MMF discontinued during SARS-CoV-2 | 2.20 (0.78-6.15) | 0.1332 | 2.00 (0.66-6.02) | 0.2174 |

\*anti-spike protein SARS-CoV-2 antibody
AKI – acute kidney injury; CI, confidence interval; cPRA, calculated panel reactive antibody; DGF, delayed graft function; DSA, donor-specific antibody; HR, hazard ratio; KDPI, Kidney Donor Profile Index; KTx, kidney transplantation; SARS-CoV-2, severe acute respiratory syndrome coronavirus 2.
Univariable and multivariable Cox proportional hazards regression analyses were performed to evaluate predictors of graft loss following SARS-CoV-2 infection. Time zero was defined as the date of the first positive SARS-CoV-2 test, and patients were followed until graft loss or administrative censoring at the end of the observation period.
Given the limited number of graft-loss events, a parsimonious adjustment strategy was prespecified. For each exposure of interest, a separate multivariable model was fitted that included the exposure together with the following baseline covariates: recipient age at transplantation, sex, donor type (deceased vs living donor), Kidney Donor Profile Index (KDPI), history of rejection, and the interval from kidney transplantation to SARS-CoV-2 diagnosis. Hazard ratios (HRs) with 95% confidence intervals (CIs) are reported. p-value < 0.05 was considered statistically significant

Model B, incorporated post-infection allograft function and immunosuppression parameters. Markers of persistent kidney injury were the strongest predictors of graft loss; higher post-SARS-CoV-2 serum creatinine (aHR 2.81, p < 0.0001), and higher albuminuria (aHR 1.09, p = 0.0007) were each strongly and independently associated with increased risk of graft-loss.

Maintenance of MMF therapy at the post-infection assessment was independently associated with a lower risk of graft failure (aHR 0.99, p = 0.0193), whereas tacrolimus exposure, and corticosteroid maintenance, were not associated with the outcome (Table 4).

**Table 4.** Univariable and multivariable SARS-CoV-2–anchored Cox proportional hazards regression analysis (Model B) of predictors of post–SARS-CoV-2 graft loss including post-infection allograft function parameters.

| Outcome: graft loss | Unadjusted HR (95% CI) | p-value | Adjusted HR (95% CI) | p-value |
| --- | --- | --- | --- | --- |
| Sex– male | 0.76 (0.35-1.61) | 0.4766 | 0.94 (0.42-2.09) | 0.8923 |
| Age at time of KTx (years) | 0.99 (0.97-1.02) | 0.9674 | 1.00 (0.97-1.04) | 0.6804 |
| Deceased donor | 1.27 (0.60-2.71) | 0.5256 | 1.26 (0.56-2.81) | 0.5666 |
| KDPI | 1.01 (0.98-1.05) | 0.2677 | 1.02 (0.98-1.06) | 0.2187 |
| History of rejection | 2.61 (1.17-5.84) | **0.0188** | 1.99 (0.56-7.02) | 0.2847 |
| Time from KTx to SARS-CoV-2 testing (months) | 1.00 (1.00-1.01) | **0.0065** | 1.01 (0.98-1.04) | 0.3048 |
| Time from SARS-CoV-2 testing to study enrollment | 1.00 (0.99-1.01) | **0.0389** | 0.99 (0.98-1.00) | 0.0554 |
| Creatinine (mg/dL) | 1.92 (1.53-2.40) | **<0.0001** | 2.81 (1.87-4.24) | **<0.0001** |
| Urine Albumin/Creatinine Ratio - UACR (mg/g) | 1.07 (1.03-1.10) | **0.0001** | 1.09 (1.03-1.14) | **0.0007** |
| C-reactive protein (mg/L) | 1.00 (0.98-1.03) | 0.5496 | 1.01 (0.98-1.05) | 0.2492 |
| Positive urine for BK virus | 0.72 (0.17-3.04) | 0.6574 | 0.76 (0.17-3.30) | 0.7156 |
| Systolic blood pressure (mm Hg) | 1.01 (0.99-1.03) | 0.1880 | 1.01 (0.99-1.04) | 0.2071 |
| Diastolic blood pressure (mm Hg) | 1.00 (0.95-1.04) | 0.9997 | 0.98 (0.93-1.04) | 0.6581 |
| Heart rate (bpm) | 1.02 (0.99-1.06) | 0.1473 | 1.02 (0.98-1.06) | 0.2193 |
| Body mass index (kg/m<sup>2</sup>) | 0.98 (0.92-1.05) | 0.7407 | 0.97 (0.90-1.05) | 0.5639 |
| DSA before COVID-19 | 1.87 (0.80-4.39) | 0.1460 | 1.61 (0.55-4.67) | 0.3760 |
| Positive anti-S* antibodies after COVID-19 | 1.48 (0.19-11.18) | 0.6984 | 1.96 (0.16-23.40) | 0.5930 |
| Tacrolimus maintenance | 0.96 (0.81-1.12) | 0.6150 | 0.96 (0.83-1.11) | 0.6324 |
| MMF maintenance | 0.9981 (0.9968-0.9994) | **0.0034** | 0.9985 (0.9973-0.9998) | **0.0193** |
| Steroid maintenance | 1.60 (0.75-3.43) | 0.2206 | 0.83 (0.28-2.46) | 0.7433 |

\*anti–spike protein SARS-CoV-2 antibody
Univariable and multivariable Cox proportional hazards regression analyses were performed to evaluate associations between post–SARS-CoV-2 allograft function, immunosuppression parameters, and subsequent graft loss. Time zero was defined as the date of the first positive SARS-CoV-2 test.
Model B used the same prespecified baseline adjustment set as Model A (recipient age at transplantation, sex, donor type, KDPI, history of rejection, and interval from kidney transplantation to SARS-CoV-2 diagnosis) and was additionally adjusted for the interval between SARS-CoV-2 diagnosis and the Time from SARS-CoV-2 testing to study enrollment at which the post-COVID parameters were recorded.
For each post-COVID parameter of interest, a separate multivariable model was constructed. Hazard ratios (HRs) with 95% confidence intervals (CIs) are reported. p-value < 0.05 was considered statistically significant.

To integrate these findings, we constructed a final multivariable model including all variables that were significant in either Model A or Model B, adjusted using the same covariate set as Model B. In this combined model, unvaccinated status, higher post-infection serum creatinine, higher albuminuria, and MMF dose remained independently associated with graft loss (Supplementary Table S10).

#### Transplantation-Anchored Long-Term Graft Survival

In transplantation-anchored Cox proportional hazards analyses (time zero at kidney transplantation), unvaccinated status was significantly associated with long-term graft loss in univariable analysis. In the prespecified multivariable model that included recipient age and sex, donor type, Kidney Donor Profile Index, history of rejection, delayed graft function, and repeat transplantation, unvaccinated status remained independently associated with a higher risk of graft loss (aHR 2.80, 95% CI 1.07–7.30, p = 0.0342). None of the other covariates retained statistical significance after adjustment (Table 5).

**Table 5.** Univariable and multivariable transplantation-anchored Cox proportional hazards regression analysis of predictors of long-term graft loss.

| Outcome: graft loss | Unadjusted HR (95% CI) | p-value | Adjusted HR (95% CI) | p-value |
| --- | --- | --- | --- | --- |
| Sex – male | 1.62 (0.68-3.83) | 0.2708 | 1.43 (0.53-3.81) | 0.4701 |
| Age at time of KTx (years) | 1.02 (0.99-1.06) | 0.0944 | 1.01 (0.97-1.05) | 0.3902 |
| Deceased donor | 1.93 (0.87-4.27) | 0.1049 | 1.91 (0.63-5.75) | 0.2453 |
| KDPI | 1.03 (0.99-1.07) | 0.0551 | 1.04 (0.99-1.11) | 0.0791 |
| History of rejection | 1.42 (0.60-3.38) | 0.4215 | 1.16 (0.37-3.56) | 0.7945 |
| Delayed graft function | 2.26 (0.88-5.78) | 0.0891 | 1.58 (0.69-8.04) | 0.4182 |
| Repeat KTx | 0.95 (0.23-3.21) | 0.9399 | 0.99 (0.14-6.97) | 0.9982 |
| Unvaccinated status | 2.83 (1.26-6.35) | **0.0113** | 2.80 (1.07-7.30) | **0.0342** |

AKI, acute kidney injury; CI, confidence interval; DGF, delayed graft function; HR, hazard ratio; KDPI, Kidney Donor Profile Index; KTx, kidney transplantation; SARS-CoV-2, severe acute respiratory syndrome coronavirus 2.
Cox proportional hazards regression analyses were performed to evaluate predictors of long-term graft loss, with time zero defined as the date of kidney transplantation. Time to event was calculated from transplantation to graft loss, and patients without graft loss were administratively censored at the end of the observation period.
The multivariable model was prespecified and included the following covariates: recipient sex, age at transplantation, donor type (deceased vs living), Kidney Donor Profile Index (KDPI), history of rejection, delayed graft function (DGF), repeat transplantation ( $\geq 2$ nd KTx), and COVID-19 vaccination status (never-vaccinated vs ever-vaccinated). Hazard ratios (HRs) with 95% confidence intervals (CIs) from univariable and multivariable analyses are presented. p-value < 0.05 was considered statistically significant.

## 4 DISCUSSION

COVID-19 has been associated with substantial acute morbidity and mortality among kidney transplant recipients [29,30,32,38]. Although vaccination has improved short-term outcomes in this population [20–31,34,39], the long-term impact of SARS-CoV-2 infection and the durability of vaccine-associated protection for allograft and patient survival remain poorly characterized [32, 38,40]. In this cohort with more than four years of follow-up after SARS-CoV-2 infection, vaccination status, AKI during infection, post-infection allograft function, and maintenance immunosuppression each emerged as key determinants of late graft outcomes.

In our study, COVID-19 vaccination was independently associated with significantly better long-term graft outcomes, suggesting a durable protective effect that extends well beyond the acute phase of infection. Interestingly, vaccination administered before SARS-CoV-2 infection did not influence long-term graft survival, whereas ever-vaccinated status - vaccination at any time before or after infection-remained strongly protective [31]. Beyond biological mechanisms, behavioral and health engagement factors may contribute: lower adherence to medical recommendations, greater mistrust in healthcare, and reduced engagement in follow-up care [41–45]. Although behavioral data were not captured in this study, such patterns may partly underlie the persistently elevated graft-loss risk observed in never-vaccinated recipients, even if their impact is likely attenuated in our cohort, in which all patients were actively followed.

AKI associated with SARS-CoV-2 infection emerged as one of the strongest predictors of subsequent graft loss. Logistic regression identified absence of vaccination, dyspnea, diarrhea, and lack of pre-existing SARS-CoV-2 antibodies as independent predictors of AKI, supporting the concept that impaired antiviral immunity and clinically severe infection predispose to renal injury. These findings align with mechanistic and clinical evidence linking COVID-19–associated AKI to tubular injury, endothelial dysfunction, microvascular instability, and cytokine-mediated damage [14,15,46–48]. Importantly, AKI remained a strong independent predictor of graft loss after adjusting for comorbidities, immunologic risk, and donor factors, indicating that AKI represents not only a marker of acute disease severity but also a biologically consequential event with lasting impact on allograft viability.

Hospitalization for COVID-19 did not independently predict long-term graft loss. Logistic regression showed that hospitalization was primarily driven by symptoms reflecting acute systemic involvement, consistent with prior reports of more pronounced acute dysfunction among hospitalized transplant recipients [5–7,49,50]. Because all patients with AKI in our cohort were hospitalized, and AKI itself was strongly associated with graft failure, the adverse outcomes often attributed to hospitalization likely reflect the development of AKI rather than hospitalization per se. In adjusted models, recipients with repeat transplants were also more likely to require hospitalization, a pattern that likely reflects closer clinical surveillance and a lower admission threshold among patients presenting with higher creatinine and lower eGFR.

Post–SARS-CoV-2 serum creatinine, and UACR were associated with graft loss, but given variable timing and absence of pre-infection baselines, these measures cannot be interpreted as COVID-specific injury. Instead, they function as prognostic indicators of underlying allograft dysfunction detectable at the time of post-infection assessment.

Management of antimetabolite therapy during COVID-19 remains an important unresolved clinical question. Early expert recommendations favored MMF reduction [16,17], whereas later reports raised concerns regarding the potential alloimmune risks of prolonged minimization. Institutional practice at NYP–Weill Cornell Medicine during the study period generally favored MMF dose reduction rather than complete discontinuation [1,2]. In our cohort, neither MMF reduction nor discontinuation during acute SARS-CoV-2 infection was associated with graft loss in the SARS-CoV-2–anchored model. By contrast, continuation of MMF at the post-infection assessment was independently associated with a lower risk of graft loss in the model incorporating post-COVID allograft function parameters. Notably, an elegant study demonstrated that MMF can inhibit SARS-CoV2 entry into a lung organoids derived from human pluripotent cells [51]. However, because MMF exposure was recorded at a single post-infection time point and treatment decisions reflected individualized clinical judgment rather than a standardized protocol, this association likely reflects greater clinical and immunologic stability in patients for whom MMF could be maintained, rather than a direct protective effect of the drug itself. Nevertheless, this observation aligns with growing evidence that prolonged or complete withdrawal of antimetabolites may increase alloimmune risk [52,53].

Taken together, vaccination status, AKI during infection, and post-infection allograft function identify kidney transplant recipients at heightened long-term risk following SARS-CoV-2 infection, although the underlying mechanisms are likely multifactorial. The protective association of vaccination -irrespective of timing relative to infection – has particular clinical relevance in the context of evolving viral variants and updated vaccine formulations.

In multivariable Cox models, never-vaccinated status and AKI associated with SARS-CoV-2 infection independently predicted late graft failure. When post-infection renal function parameters were included, serum creatinine and albuminuria were the strongest predictors of subsequent graft loss, whereas higher maintenance MMF dose was associated with a lower risk of graft failure.

A potential limitation of this study is that it a single-center design. However, single center studies offer several advantages over multicenter studies including greater consistent in protocols and procedures, uniform data quality, better control over confounders and strong internal validity. Post-infection assessments were performed at variable time points, which may introduce some heterogeneity; however all measurements were obtained during routine clinical follow-up. Immunosuppression data, including MMF dosing, were captured at a single post-infection time point., reflecting standard practice rather than dynamic changes. The number of AKI events was relatively small which may limit statistical precision, although key associations remained robust in multivariable models. Despite these limitations, the study has notable strengths: it represents the longest follow-up cohorts of kidney transplant recipients with confirmed SARS-CoV-2 infection; it employs complementary transplantation-anchored and SARS-CoV-2-anchored Cox models; and it includes detailed clinical, virologic, immunologic, and transplant-related variables.

In this long-term cohort, COVID-19 vaccination emerged as a major determinant of favorable post-infection outcomes, whereas AKI identified a particularly high-risk subgroup with persistently elevated graft-failure risk.

## 5 CONCLUSION

COVID-19 continues to exert long-term effects on kidney transplant recipients. In this cohort, unvaccinated recipients experienced significantly worse graft survival during the four years following SARS-CoV-2 infection, underscoring the importance of sustained antiviral immune protection. AKI during COVID-19 identified a particularly high-risk subgroup with persistently elevated graft failure risk, while post-infection allograft function provided additional prognostic insight. Patterns of immunosuppression adjustment varied; however, continuation of MMF appeared beneficial.

As SARS-CoV-2 variants evolve and vaccine formulations adapt, these findings reinforce a central message: maintaining COVID-19 vaccination in kidney transplant recipients remains a critical strategy for long-term graft preservation and patient protection.

## Acknowledgments

We thank Gabriel Stryjniak for REDCap programming and assistance with data collection. We are deeply grateful to the patients who participated in this study and to the entire clinical and research team involved in their care and in the conduct of this project.

## Data availability statement

Anonymized data that support the findings of this study are available from the corresponding author upon reasonable request, subject to approval by the local ethics committee and data protection regulations.

## Supporting information statement

Additional supporting information may be found online in the Supporting Information section.

## Author contributions

I.D. - conception of the analysis, data curation, statistical analysis, interpretation of results, drafting of the manuscript, incorporation of coauthor revisions, and preparation of the final version.

D.M.D. - design and writing of the clinical IRB protocol, design of REDCap database, enrollment and consenting of patients, data validation, and review and editing of the abstract and manuscript.

C.L. - laboratory database management of enrolled patients

N.H. - recruitment and clinical management of patients

P.L. - initial data collection, initial analysis, and abstract presentation of the data.

J.R.L. - recruitment and clinical management of patients.

M.T. - recruitment and clinical management of patients.

M.S. - overall study design, supervision, and critical review and editing of the abstract and manuscript.

## Abbreviations

AKI: acute kidney injury
ATG: antithymocyte globulin
BMI: body mass index
COVID-19: coronavirus disease 2019
CRP: C-reactive protein
cPRA: calculated panel reactive antibody
DBD: donor after brain death
DCD: donation after circulatory death
DGF: delayed graft function
DSA: increased donor-specific antibody
ECD: expanded criteria donor
eGFR: estimated glomerular filtration rate
KDPI: Kidney Donor Profile Index
MFI: mean fluorescence intensity
MMF: mycophenolate mofetil
NYP-WCM: New York Presbyterian Hospital-Weill Cornell Medicine
PCR: polymerase chain reaction
SARS-CoV-2: Severe Acute Respiratory Syndrome Coronavirus 2
SCD: standard criteria donor
UACR: urine albumin-to-creatinine ratio

